# Indirect effects of peaks in COVID admissions on access to surgery in the English NHS, differential effects by operation type, ethnicity and socio-economic status: a database study

**DOI:** 10.1101/2021.09.09.21262542

**Authors:** Sandra C Remsing, Felicity Evison, Ravinder S Vohra, Dion Morton, Peter Chilton, Kamlesh Khunti, Richard J Lilford

**Affiliations:** Department of Health Informatics, University Hospitals Birmingham NHS Foundation Trust, Edgbaston, Birmingham, UK; Trent Oesophago-Gastric Unit, Nottingham City Hospital, Nottingham, UK; Institute of Cancer and Genomic Sciences, University of Birmingham, Edgbaston, Birmingham UK; Institute of Applied Health Research, University of Birmingham, Edgbaston, Birmingham UK; Diabetes Research Centre, University of Leicester, Leicester, UK

**Keywords:** COVID, Hospital admission, Surgery

## Abstract

**Objectives:** During the COVID pandemic the UK saw two peaks in the prevalence of hospital admissions resulting in disruption of routine hospital services in the English National Health Service. This study aimed to track the effect of these peaks on various types of surgery representing differences in urgency, importance, and complexity.

**Design:** Database study using the Hospital Episode Statistics database and surgical operations selected purposively, to represent different combinations of urgency, importance and complexity.

**Setting:** All hospitals within England that carried out procedures funded by the National Health Service.

**Main Outcome Measures:** Number of emergency routine surgeries; cancer-removal surgeries; transplant surgeries; renal transplants Deceased and living donors); and elective routine surgeries carried out prior to and during the COVID pandemic.

**Results:** While all surgeries declined, emergency or urgent operations held up better than elective cases. There was rapid rebound between peaks. Among emergency cases, coronary angioplasty for acute myocardial infarction held up well in contrast to appendectomy, where indications for surgery are more elastic. Among urgent cancer and transplant operations, those with the most complex pathways were the most severely affected. The pandemic affected socio-economic and ethnic groups similarly. Disruption during the second peak was slightly less than during the first peak despite even greater COVID admission rates.

**Conclusion:** The NHS titrated its response appropriately to the pandemic by prioritising emergency and urgent cases over elective cases. However, complex time critical conditions like organ transplants and certain cancers are also disrupted with implications for third peaks in hospital admissions that many countries are experiencing.

## Introduction

The UK has been severely affected by the COVID pandemic, in particular over two peaks in prevalence.^1^ The English National Health Service (NHS) has been unable to maintain routine services at their previous levels,^2,3^ and in certain instances operating theatre capacity was used to manage an overspill from intensive care units.^4^ One manifestation of this stress on the service has been a build-up in waiting lists for interventional procedures,^5^ henceforth described simply as ‘surgery’. During the first peak, a number of national organisations (including NHS England, NHS Blood and Transplant, Royal College of Surgeons, NICE) issued guidelines on service priorities. On the 20/03/2020 the Royal Colleges of Surgeons advised its members to prioritise emergency services.^6^ In this paper, we track the use of purposively selected types of surgical procedure over two peaks in the pandemic. First, we examine the effect of the pandemic by type of surgery. Second, we examine for any differential effect on use of the selected surgeries according to social class and ethnic group. Third, we examine for any effect of peaks in the pandemic on readmission rates and mortality associated with the selected surgeries.

The surgeries were selected to shed light on three factors that are likely to determine access to surgery during a pandemic.^7,8^ First, degree of urgency, favouring emergency procedures. Second, potential for benefit, favouring life-saving procedures. Third, degree of complexity, favouring less complex surgeries. We acknowledge that these factors are correlated and therefore can be only partially disentangled, a point to which we return in the discussion.

## Methods

Data were extracted from the Hospital Episodes Statistics (HES) database that contains all emergency and elective patient admissions, outpatient appointments and A&E attendances funded by the NHS in England. All procedures funded by the NHS were included, irrespective of whether they took place in NHS or independent hospitals. Our study was based on adult patients (aged ≥18 years old) with the selected operation codes.

For emergency procedures, we selected appendectomy and coronary angioplasty for ST-Elevation Myocardial Infarction (STEMI). For non-emergency conditions that are nevertheless lifesaving, we selected primary surgical procedures for cancer. To examine complexity we took a number of approaches. First, we selected cancer diagnoses in two categories: those where the care pathway is relatively straightforward (breast lumpectomy, hemi-colectomy) versus those where more complex pathways (that may include intensive) are required (pancreatectomy, hemi-hepatectomy and eosphagectomy). Second, we examined transplant surgery. Our main reason for selection this class of procedures was to compare renal transplants from living versus deceased donors. Living donor transplants are particularly complex involving the co-ordination of surgical procedures in many different parts of the country on a single day. Lastly, we selected two ‘control’ conditions to represent common elective procedures of no particular urgency: hip replacement and primary inguinal hernia repair.

We extracted data from admissions recorded between January 2015 and March 2021 (the specific terms used and their codes are given, alongside guidance issued in accordance to each code, in Table 1). Patients who died within 90 days of discharge were identified, as were patients who had an emergency readmission for any reason within 90 days. For the cohort of patients undergoing renal transplantation, the donor type was extracted to compare outcomes in those receiving an organ from a live donor versus a deceased donor. Length of hospital stay (LOS) was calculated from admission date to discharge date (or hospital death date) for each spell. Patients with missing information sex, ethnicity or age were excluded, as well as patients with inconsistent records such as having subsequent admissions after death date. (Note that missing ethnicity refers to patients where no information on ethnicity is provided. This is not the same as ‘unknown’ ethnicity, which is coded for in HES (‘Z’, ‘99’) and is therefore included in the analysis.) We created a series of graphs to show how the monthly procedure rates changed in relation to the peaks in the pandemic. The peaks were defined from the date of complete UK lockdown (26^th^ March 2020 – 3 July 2020, 5^th^ November 2020 to 2^nd^ December 2020, 6^th^ January 2021 – ongoing up to 31 March 2021).

**Table 1:**
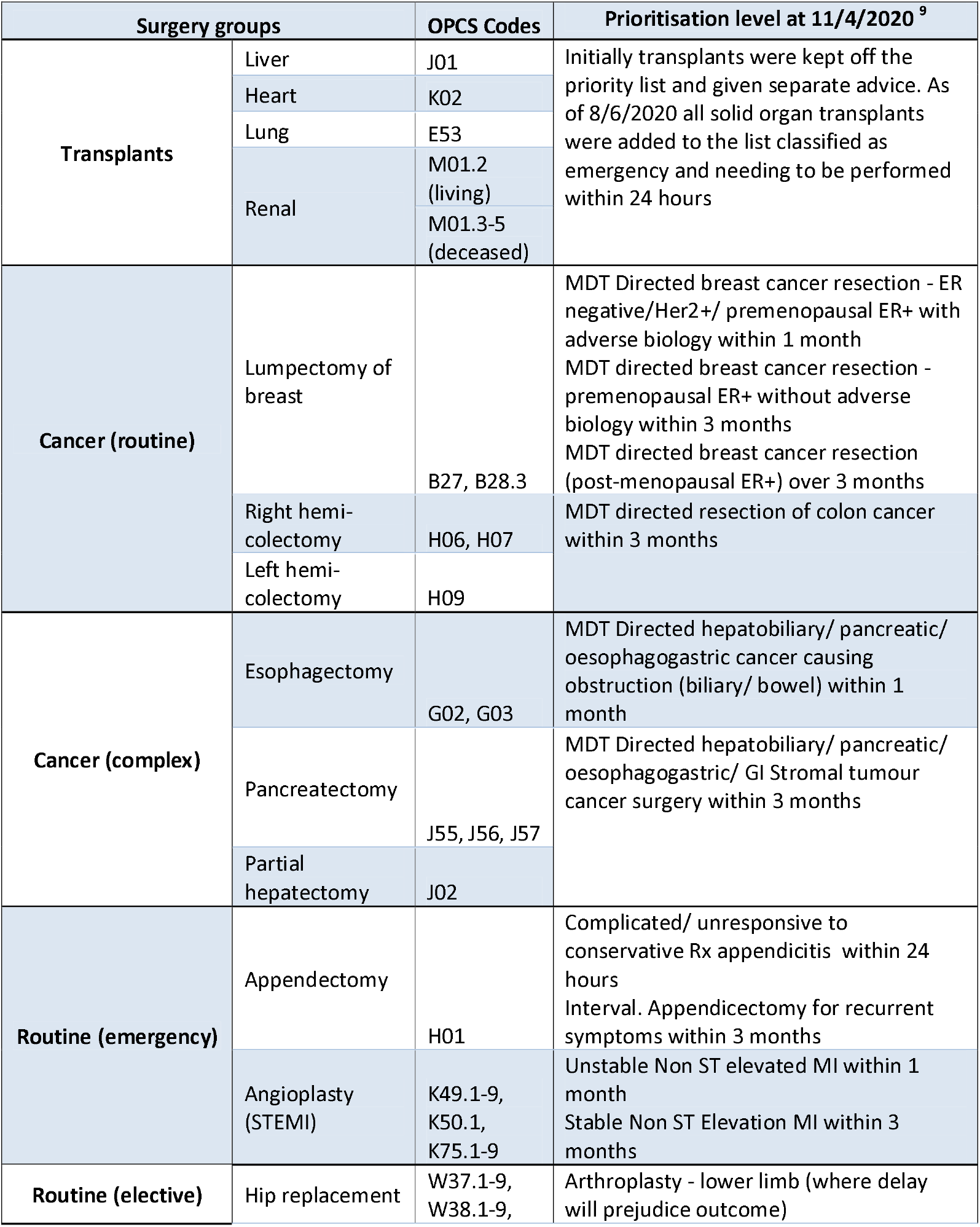
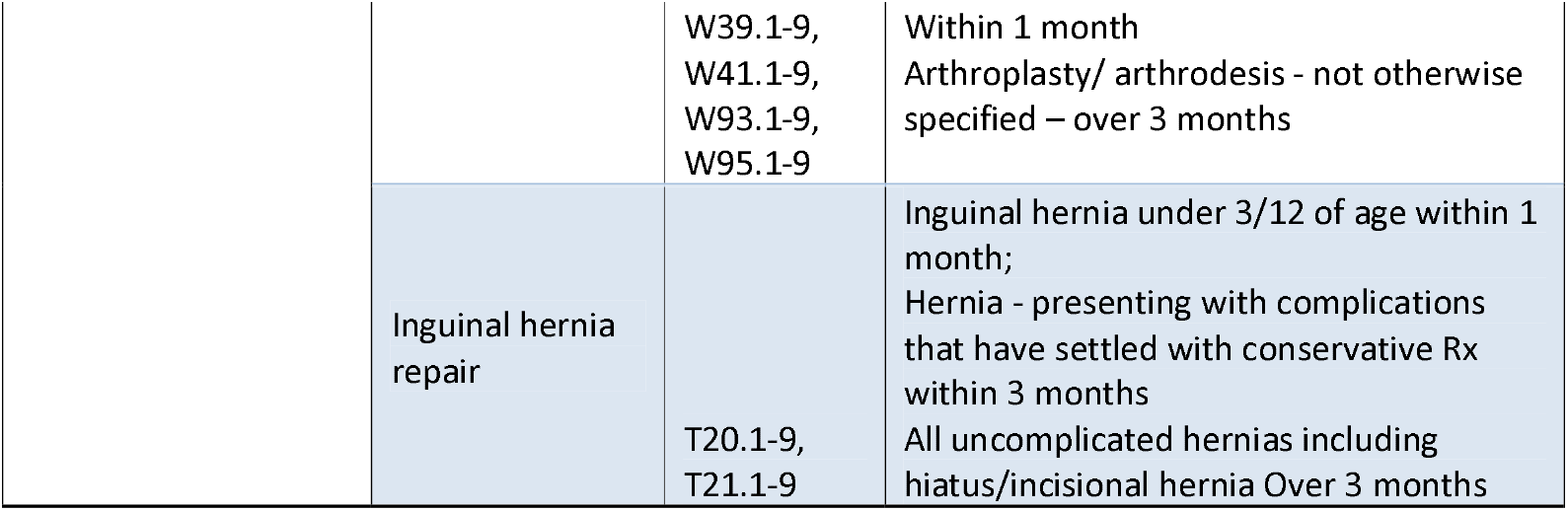
Surgery groups and operation codes

### Statistical analysis

The graphical representations track admissions from January 2015 to the end of March 2021, the latest figures released by NHS Digital. This covers both peaks of COVID admissions. For the purposes of statistical analysis, we do not include the period to March 2021 because we wish to include the follow-up time for 90-day deaths and readmissions. Our statistical analysis is therefore based on a 6-month period (Mar-Aug) for 2015-2019, and for 2020. This is therefore not the same as the figures, which plot the number of surgeries from Jan 2015 – March 2021. We have isolated our analysis to this 6-month window Mar-Aug to coincide with the first wave of the COVID pandemic. The analysis therefore provides a useful snapshot into what happened in the first COVID peak compared to the same time of year in the five previous years.

The total number of surgeries were counted and grouped by type, ethnicity, deprivation and sex, and split, as stated, into two comparative time-periods (March-August 2015-2019 and March-August, 2020). Chi-squared tests were performed to compare the proportions of surgeries occurring between these groups and time-periods. Similarly, the total number of emergency readmissions and deaths within 90 days were also counted and grouped by period and compared by chi-squared tests. Median age and median LOS were calculated and we used a Mann-Whitney U test to compare the distribution between the two time-periods. Additionally, we reported the relative difference in the number of surgeries between time-periods as an average six-month percent change. We calculated this by taking the average number of operations per year in the pre-COVID time window (total divided by 5 years) and comparing this to the total number of operations in the corresponding time-period in 2020. A percentage difference between the two was then calculated and reported as the percentage difference in 2020 comparatively (‘plus’ or ‘minus’ sign indicating direction).

Removal surgeries were then paired with corresponding relevant primary cancer-diagnoses codes to isolate cancer-specific removal surgeries. Cancer codes B50 (breast cancer), C18-C20 (colon and colorectal cancer), C25 (pancreatic cancer), C22 (liver cancer) and C15 (oesophageal cancer) were used. Similarly, angioplasty surgeries were paired with a primary diagnosis of STEMI (I21.0-3).

## Results

We identified 1,184,192 eligible procedures that occurred across England between 1 January 2015 and 31 March 2021. Of these, 20,752 were transplants, 113,134 were cancer-removal surgeries (complex and routine) and 1,050,306 were routine procedures (elective and emergency). Figures 1-4 show a substantial reduction in the number of surgeries taking place from March/April 2020 corresponding to the first peak in COVID. Perhaps most notable is the sudden dip in the number of emergency appendectomies in March 2020; a pattern not mirrored for emergency angioplasties (Figure 1). Cancer-removal surgeries were also severely disrupted during the first lockdown, with a visible drop in the number of surgeries occurring from March 2020 across both routine and complex surgeries. During the second peak, however, less complex cancer surgery held up better than the more complex cases (Figure 2). Transplants show a similar pattern to complex cancer cases, with a notable drop coinciding with the first lockdown, relatively quick recovery, only to be severely disrupted again with subsequent lockdowns (Figure 3). The most prominent disruption is seen with renal transplants from living donors, which reduced to zero for a couple of months as a direct result of the March 2020 lockdown. These transplants pick up again in the months to follow, although not to the levels recorded prior to the pandemic. Elective surgery (Figure 4) declined almost to zero in the first COVID peak, but held up rather better in the second.

**Figure 1:**
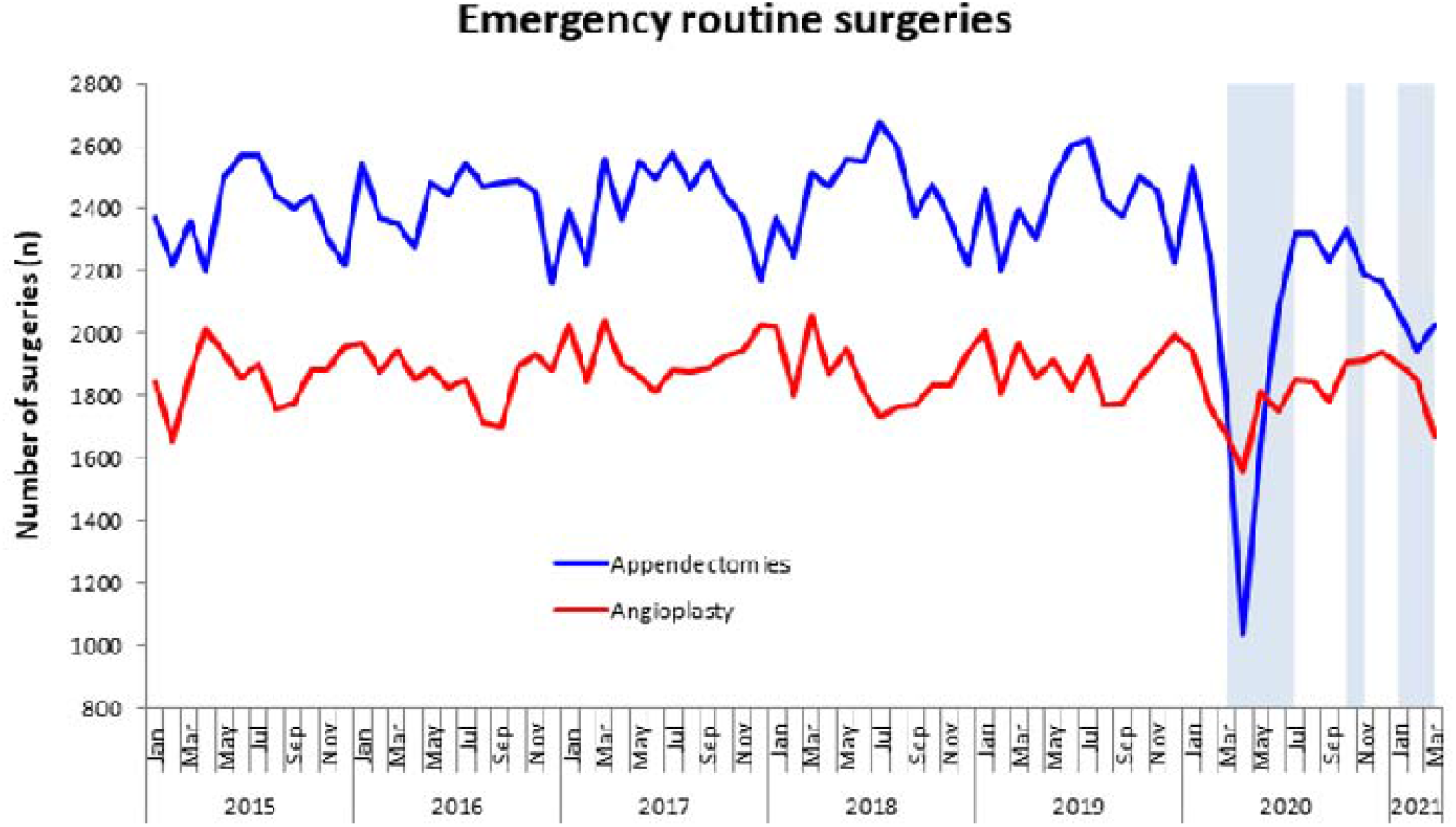
Number of emergency routine surgeries, January 2015 – March 2021. *Note*. UK National COVID Lockdowns denoted by shaded regions.

**Figure 2:**
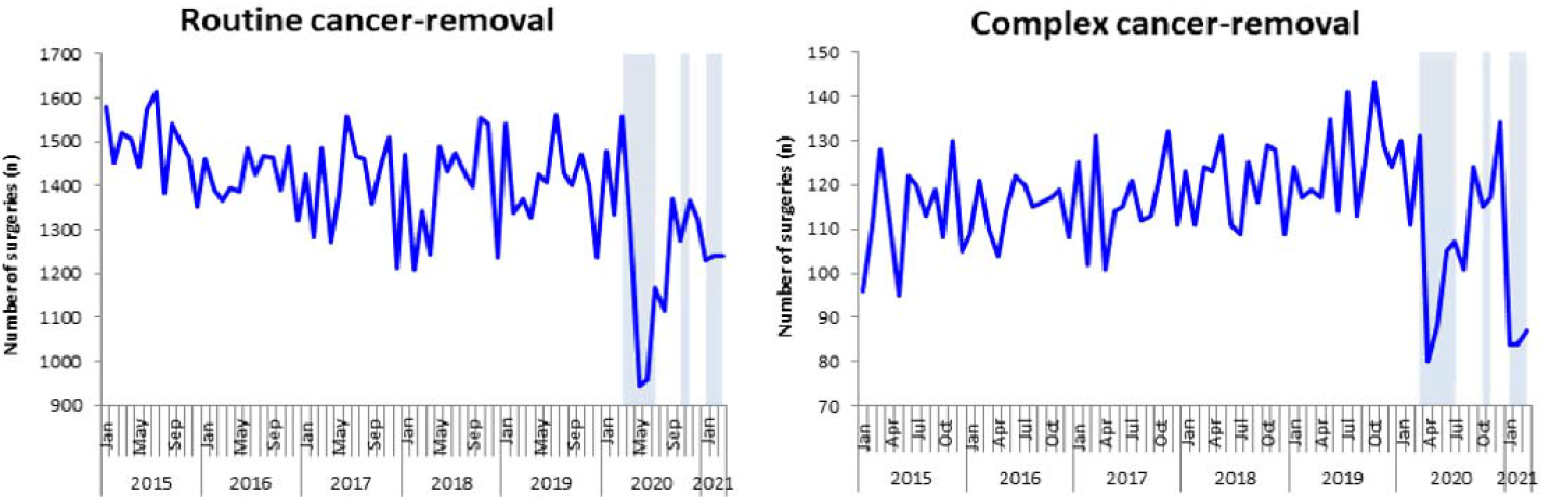
Number of cancer-removal surgeries, routine vs complex, January 2015 - March 2021. *Note*. UK National COVID Lockdowns denoted by shaded regions.

**Figure 3:**
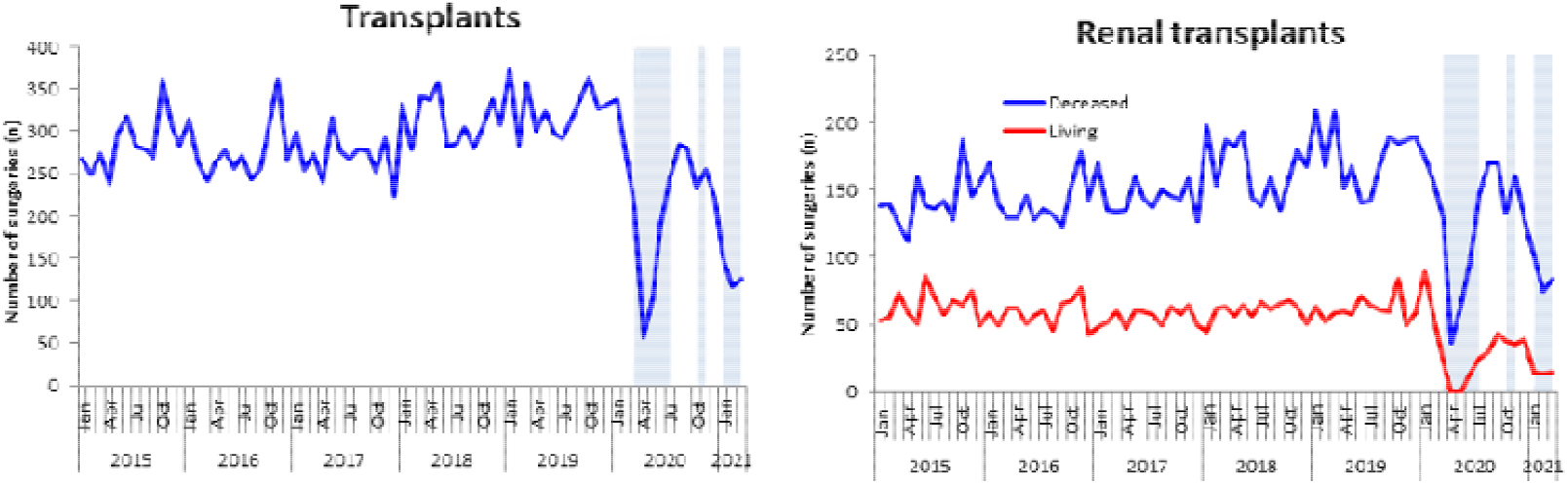
Number of total transplant surgeries, and renal transplants, January 2015 to March 2021. *Note*. UK National COVID Lockdowns denoted by shaded regions.

**Figure 4:**
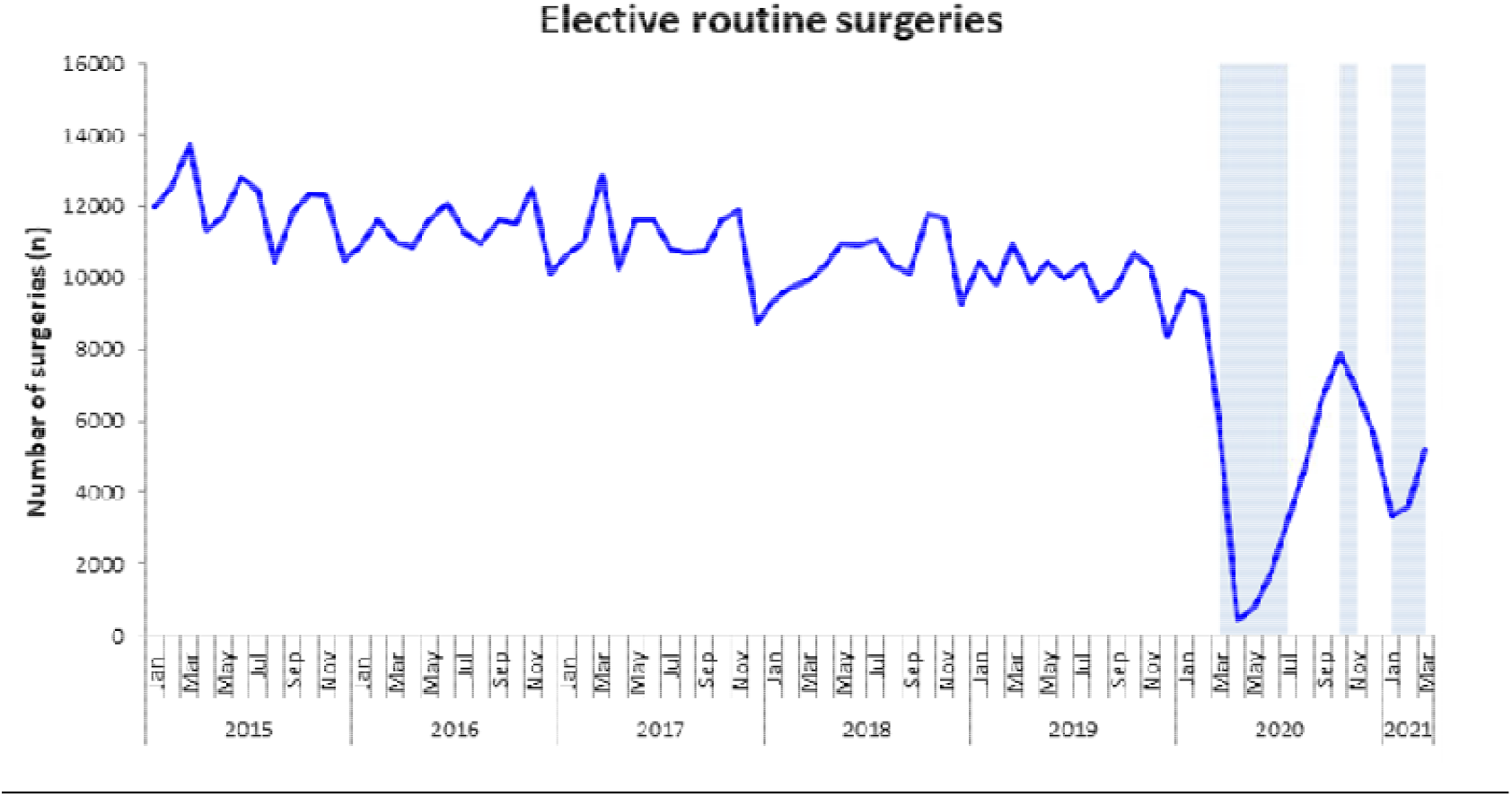
Number of elective routine surgeries, January 2015 to March 2021. *Note*. UK National COVID Lockdowns denoted by shaded regions.

A closer look at the direct comparison between the ‘COVID’ first wave time window (March – August 2020) and the equivalent time window in prior years, ‘pre-COVID’ (March-August, 2015 -2019) shows an average six-month percentage decrease across all surgeries and demographic groups (Tables 2-5). Noteworthy is the observed percentage reduction in the number of renal transplants from living donors in 2020 compared to those in the previous five years; a finding likely impacted by the total cessation of these transplants for two months during the first lockdown (29% pre-COVID to 12% during COVID) (Table 4). Similarly, notable decreases in average six-month percent change are seen in inguinal hernia repairs and hip replacements (72% and 77% reductions in 2020 respectively), while emergency appendectomies only decrease by 25% on average, possibly due to the quick recovery after the peak of the pandemic (Table 2 and 5).

**Table 2:** Emergency surgeries summary statistics comparing two time windows: Pre-COVID (March – August, 2015 – 2019), COVID (March-August, 2020).

|  | **Pre-covid**<br>**(2015-2019)** | **Covid (2020)** | **p-value** | **Avg (n) per year (pre-covid**<br>**total/5yrs)** | **avg annual % change (Covid**<br>**compared to avg pre_covid)** |
| --- | --- | --- | --- | --- | --- |
| **Emergency routine surgeries (n)** | 130695 | 21718 | **<0.001** | 26139 | -17% |
| **Type** |  |  |  |  |  |
| Angioplasties (STEMI) | 56228 (43%) | 10506 (48%) |  | 11246 | -7% |
| Appendectomy | 74467 (57%) | 11212 (52%) | **<0.001** | 14893 | -25% |
| **Sex (n)** |  |  |  |  |  |
| Male | 78884 (60%) | 13644 (63%) |  | 15777 | -14% |
| Female | 51811 (40%) | 8074 (37%) | **<0.001** | 10362 | -22% |
| **Ethnicity (n)** |  |  |  |  |  |
| White | 98812 (75%) | 15844 (73%) |  | 19762 | -20% |
| South Asian | 7223 (6%) | 1130 (5%) |  | 1445 | -22% |
| Black | 2245 (2%) | 342 (2%) |  | 449 | -24% |
| Mixed/other | 3793 (3%) | 647 (3%) |  | 759 | -15% |
| Unknown | 18622 (14%) | 3755 (17%) | **<0.001** | 3724 | (+) 1% |
| **Deprivation (n)** |  |  |  |  |  |
| Quintile 1 (most deprived) | 26560 (20%) | 4282 (20%) |  | 5312 | -19% |
| Quintile 2 | 26570 (20%) | 4167 (19%) |  | 5314 | -22% |
| Quintile 3 | 25449 (20%) | 4303 (20%) |  | 5090 | -15% |
| Quintile 4 | 24342 (19%) | 4148 (19%) |  | 4868 | -15% |
| Quintile 5 (most affluent) | 23052 (18%) | 3845 (18%) |  | 4610 | -17% |
| Unknown | 4722 (3%) | 973 (4%) | **<0.001** | 944 | (+) 3% |
| **Age (years) (median, IQR)** | 50 (31 to 65) | 53 (35 to 56) | **<0.001** | n/a | n/a |
| **LOS (days) (median, IQR)** | 3 (2 to 4) | 2 (2 to 4) | **<0.001** | n/a | n/a |
| **Deaths from any cause (within 90 days) (n)** | 4028 (3%) | 770 (4%) | **<0.001** | n/a | n/a |
| **Emergency readmissions (within 90 days) (n)** | 15509 (12%) | 2789 (13%) | **<0.001** | n/a | n/a |

\*p-value shows whether there is a statistically significant difference in the compositional makeup of patients within each subgroup, and between the two time periods.

**Table 3:** Cancer-removal summary statistics comparing two time windows: Pre-COVID (March – August, 2015 – 2019), COVID (March-August, 2020).

|  | Pre-covid (2015-2019) | Covid (2020) | p-value | Avg (n) per year (pre-covid total/5yrs) | avg annual % change (Covid compared to avg pre-covid) |
| --- | --- | --- | --- | --- | --- |
| **Cancer removal surgeries (n)** | 46762 | 7681 | **<0.001** | 9352 | -18% |
| **Type (n)** |  |  |  |  |  |
| Partial hepatectomy | 984 (2%) | 165 (2%) |  | 197 | -16% |
| Partial pancreatectomy | 2000 (4%) | 368 (5%) |  | 400 | -8% |
| Oesophagectomy | 555 (1%) | 66 (1%) |  | 111 | -41% |
| Lumpectomy of the breast | 37133 (80%) | 6085 (79%) |  | 7426 | -18% |
| Right and left hemi-colectomy | 6090 (13%) | 997 (13%) | **0.04** | 1218 | -18% |
| **Surgery type (n)** |  |  |  |  |  |
| Routine | 43223 (92%) | 7082 (92%) |  | 8645 | -18% |
| Complex | 3539 (8%) | 599 (8%) | 0.5 | 708 | -15% |
| **Sex (%)** |  |  |  |  |  |
| Male | 5953 (13%) | 1023 (13%) |  | 1191 | -14% |
| Female | 40802 (87%) | 6658 (87%) | 0.14 | 8160 | -18% |
| **Ethnicity (%)** |  |  |  |  |  |
| White | 38054 (82%) | 5346 (70%) |  | 7611 | -30% |
| South Asian | 1534 (3%) | 258 (4%) |  | 307 | -16% |
| Black | 1016 (2%) | 162 (2%) |  | 203 | -20% |
| Mixed/other | 930 (2%) | 190 (2%) |  | 186 | (+)2% |
| Unknown | 5228 (11%) | 1725 (22%) | **<0.001** | 1046 | (+)40% |
| **Deprivation %** |  |  |  |  |  |
| Quintile 1 (most deprived) | 7107 (15%) | 1043 (14%) |  | 1421 | -27% |
| Quintile 2 | 8196 (18%) | 1296 (17%) |  | 1639 | -20% |
| Quintile 3 | 9610 (21%) | 1555 (20%) |  | 1922 | -19% |
| Quintile 4 | 10303 (22%) | 1681 (22%) |  | 2061 | -18% |
| Quintile 5 (most affluent) | 10419 (22%) | 1768 (23%) |  | 2084 | -15% |
| Unknown | 1127 (2%) | 338 (4%) | **<0.001** | 225 | (+)33% |
| **Age (years) (median, IQR)** | 64 (51 to 74) | 62 (50 to 73) | **<0.001** | n/a | n/a |
| **LOS (days) (median, IQR)** | 1 (1 to 5) | 1 (0 to 3) | **<0.001** | n/a | n/a |
| **Deaths from any cause (within 90 days) (n)** | 597 (1%) | 83 (1%) | 0.15 | n/a | n/a |
| **Emergency readmissions (within 90 days) (n)** | 4869 (10%) | 676 (9%) | **<0.001** | n/a | n/a |

\*p-value shows whether there is a statistically significant difference in the compositional makeup of patients within each subgroup, and between the two time period

**Table 4:**
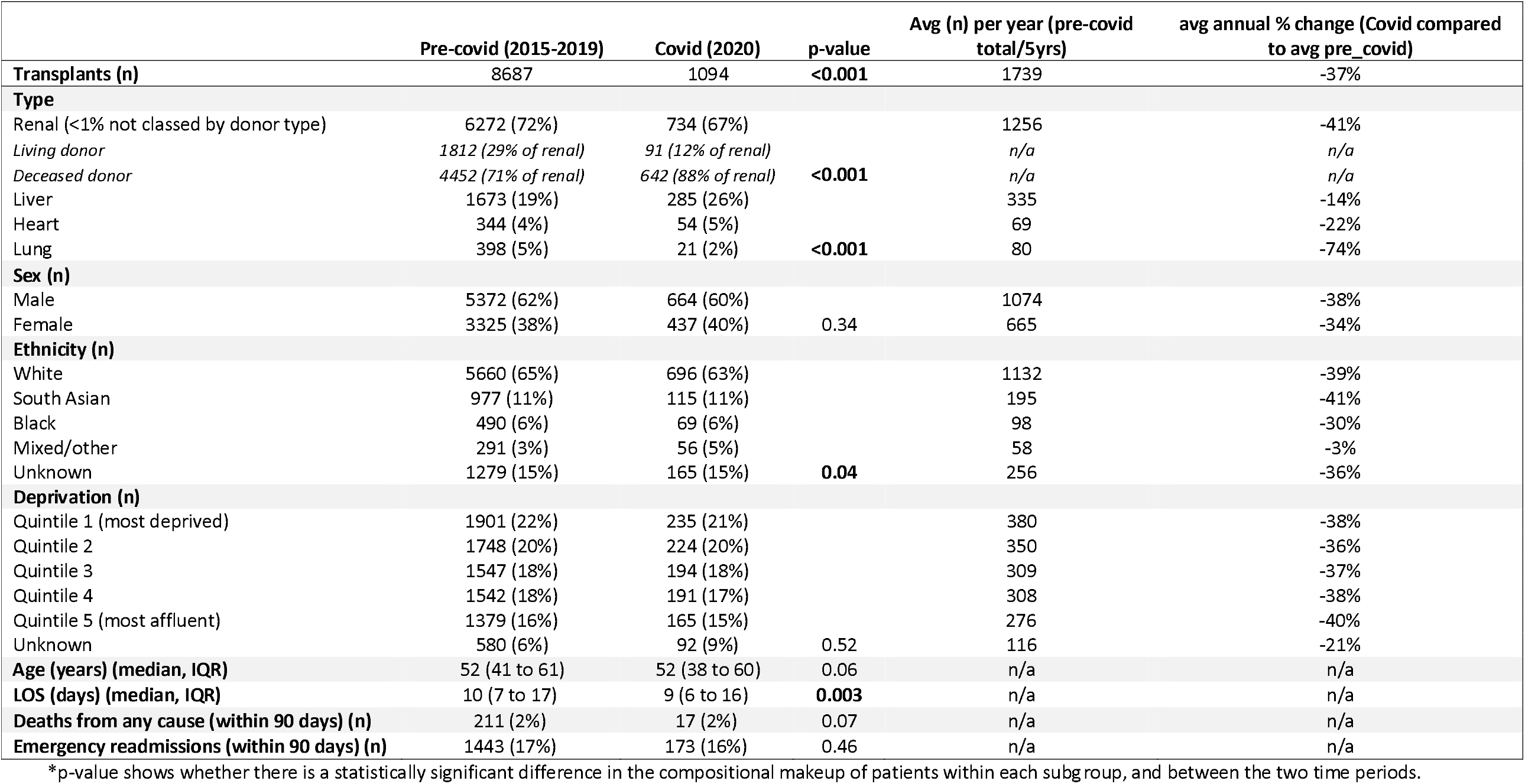
Transplants summary statistics comparing two time windows: Pre-COVID (March – August, 2015 – 2019), COVID (March-August, 2020).

|  | Pre-covid (2015-2019) | Covid (2020) | p-value | Avg (n) per year (pre-covid total/5yrs) | avg annual % change (Covid compared to avg pre_covid) |
| --- | --- | --- | --- | --- | --- |
| **Transplants (n)** | 8687 | 1094 | **<0.001** | 1739 | -37% |
| **Type** |  |  |  |  |  |
| Renal (<1% not classed by donor type) | 6272 (72%) | 734 (67%) |  | 1256 | -41% |
| Living donor | 1812 (29% of renal) | 91 (12% of renal) |  | n/a | n/a |
| Deceased donor | 4452 (71% of renal) | 642 (88% of renal) | **<0.001** | n/a | n/a |
| Liver | 1673 (19%) | 285 (26%) |  | 335 | -14% |
| Heart | 344 (4%) | 54 (5%) |  | 69 | -22% |
| Lung | 398 (5%) | 21 (2%) | **<0.001** | 80 | -74% |
| **Sex (n)** |  |  |  |  |  |
| Male | 5372 (62%) | 664 (60%) |  | 1074 | -38% |
| Female | 3325 (38%) | 437 (40%) | 0.34 | 665 | -34% |
| **Ethnicity (n)** |  |  |  |  |  |
| White | 5660 (65%) | 696 (63%) |  | 1132 | -39% |
| South Asian | 977 (11%) | 115 (11%) |  | 195 | -41% |
| Black | 490 (6%) | 69 (6%) |  | 98 | -30% |
| Mixed/other | 291 (3%) | 56 (5%) |  | 58 | -3% |
| Unknown | 1279 (15%) | 165 (15%) | **0.04** | 256 | -36% |
| **Deprivation (n)** |  |  |  |  |  |
| Quintile 1 (most deprived) | 1901 (22%) | 235 (21%) |  | 380 | -38% |
| Quintile 2 | 1748 (20%) | 224 (20%) |  | 350 | -36% |
| Quintile 3 | 1547 (18%) | 194 (18%) |  | 309 | -37% |
| Quintile 4 | 1542 (18%) | 191 (17%) |  | 308 | -38% |
| Quintile 5 (most affluent) | 1379 (16%) | 165 (15%) |  | 276 | -40% |
| Unknown | 580 (6%) | 92 (9%) | 0.52 | 116 | -21% |
| **Age (years) (median, IQR)** | 52 (41 to 61) | 52 (38 to 60) | 0.06 | n/a | n/a |
| **LOS (days) (median, IQR)** | 10 (7 to 17) | 9 (6 to 16) | **0.003** | n/a | n/a |
| **Deaths from any cause (within 90 days) (n)** | 211 (2%) | 17 (2%) | 0.07 | n/a | n/a |
| **Emergency readmissions (within 90 days) (n)** | 1443 (17%) | 173 (16%) | 0.46 | n/a | n/a |

\*p-value shows whether there is a statistically significant difference in the compositional makeup of patients within each subgroup, and between the two time periods.

**Table 5:** Elective routine summary statistics comparing two time windows: Pre-COVID (March – August, 2015 – 2019), COVID (March-August, 2020).

|  | Pre-covid (2015-2019) | Covid (2020) | p-value | Avg (n) per year (pre-covid total/5yrs) | avg annual % change (Covid compared to avg pre-covid) |
| --- | --- | --- | --- | --- | --- |
| **Elective routine surgeries (n)** | 332913 | 17016 | <0.001 | 66583 | -74% |
| **Type** |  |  |  |  |  |
| Inguinal hernia repair | 167472 (50%) | 9397 (55%) |  | 33494 | -72% |
| Hip replacement | 165441 (50%) | 7619 (45%) | <0.001 | 33088 | -77% |
| **Sex (n)** |  |  |  |  |  |
| Male | 218920 (66%) | 11449 (67%) |  | 43784 | -74% |
| Female | 113993 (34%) | 5567 (33%) | <0.001 | 22799 | -76% |
| **Ethnicity (n)** |  |  |  |  |  |
| White | 266829 (80%) | 12834 (75%) |  | 53363 | -76% |
| South Asian | 7441 (2%) | 369 (2%) |  | 1488 | -75% |
| Black | 4013 (1%) | 192 (1%) |  | 803 | -76% |
| Mixed/other | 3826 (1%) | 233 (1%) |  | 765 | -70% |
| Unknown | 50804 (15%) | 3388 (20%) | <0.001 | 10161 | -67% |
| **Deprivation (n)** |  |  |  |  |  |
| Quintile 1 (most deprived) | 49183 (15%) | 2366 (14%) |  | 9837 | -76% |
| Quintile 2 | 59119 (18%) | 3005 (17%) |  | 11824 | -75% |
| Quintile 3 | 69688 (21%) | 3575 (21%) |  | 13938 | -74% |
| Quintile 4 | 74666 (22%) | 3758 (22%) |  | 14933 | -75% |
| Quintile 5 (most affluent) | 72751 (22%) | 3715 (22%) |  | 14550 | -74% |
| Unknown | 7506 (2%) | 597 (4%) | <0.001 | 1501 | -60% |
| **Age (years) (median, IQR)** | 66 (55 to 75) | 67 (55 to 75) | <0.001 | n/a | n/a |
| **LOS (days) (median, IQR)** | 1 (0 to 3) | 1 (0 to 3) | 0.317 | n/a | n/a |
| **Deaths from any cause (within 90 days) (n)** | 1511 (0.5%) | 186 (1%) | <0.001 | n/a | n/a |
| **Emergency readmissions (within 90 days) (n)** | 18031 (5%) | 1125 (7%) | <0.001 | n/a | n/a |

\*p-value shows whether there is a statistically significant difference in the compositional makeup of patients within each subgroup, and between the two time periods.

Ethnicity breakdowns appear consistent within patients undergoing transplants, with more differences seen in those undergoing routine surgeries. A 5% reduction in White patients undergoing routine surgeries is evident in 2020; however, this could be accounted for in part by the 3% increase in ‘Unknown’ ethnicity (Table 5). An even greater difference is seen in the percentage of ‘White’ and ‘Unknown’ patients undergoing cancer-removal surgeries, with a 12% reduction in White patients in 2020, again possibly related corresponding 11% increase in patients of Unknown ethnicity (Table 3).

Surgery outcomes remain consistent for patients undergoing cancer-removal surgeries or transplants, with little to no difference in the percentage of patient deaths or emergency readmissions within 90 days between 2015-2019 and 2020. However, a twofold increase in the percentage of deaths within 90 days is evident in 2020 for patients undergoing elective routine surgeries (0.5% to 1%, respectively, Table 5). Similarly, 90-day emergency readmissions increase, although only slightly from 5% in 2015-2019, to 7% in 2020 (see Discussion). There is an increase in deaths following emergency operations (Table 2), but this is driven by appendectomy. During the pre-COVID window 0.2% died within 90 days versus 0.4% in the COVID time-period (see Discussion).

## Discussion

### Principal findings and explanations

With the exception of coronary angioplasty, there was a sharp drop in all the selected procedures over the two peaks of COVID incidence and there was no compensatory increase in intervention rates in the period intervening between the two peaks. The prognosis for solid organ cancer is critically dependent on treatment delay,^10^ and so it must be assumed that the collateral effect of the COVID pandemic on time sensitive surgically treatable conditions was material. However, while there were sharp drops in the number of our selected procedures there were features in the data that speak to resilience in the system. The drops in incidence, while often quite dramatic, rebounded rapidly, albeit incompletely. The six-month decrease in procedure rates never exceeded 40% (observed in the case of oesophagostomy). The drop in oesophagostomy rates was likely particularly severe because of the perceived need for ongoing artificial ventilation after this operation. Coronary angioplasties did not drop appreciably despite the complexity of this procedure and the presence of multiple potential bottlenecks in the ambulance and hospital services. Our results here mirror findings previously published with respect to the first COVID peak.11,12 The data regarding emergency coronary angioplasties contrasts with the data for appendectomy where the drop in procedures was precipitous, especially in the first COVID peak. We think this might be because there is a degree of discretion regarding treatment of patients who are not critically ill or where the diagnosis is uncertain. The increased death rate for appendectomy over the COVID peak is consistent with the hypothesis that the most high-risk cases were selected for surgery during peaks in the pandemic. The increased death rate observed for elective surgery could be the result of selection effects, pandemic effects on service quality or conceivably some more direct effect on the pandemic. Deceased donor transplants dropped precipitously over both COVID peaks. Living donor kidney transplants ceased entirely, consistent with the hypothesis that the complex supply chain for living donor transplantation was severely disrupted by the pandemic.

Generally, the second peak was less disruptive than the former. The evidence does not support the explanation that it was less severe; the second peak saw, on average, a third more admissions than the first peak.^13^ We speculate that the service was able to cope better during the second peak because of lessons learned during the first peak. It is also noteworthy that advice regarding transplants changed in favour of the operation (Figure 1) but a smaller drop in incidence was seen for other operation types where no such advice had been issued.

With respect to readmissions and deaths, there are a number of data points showing significant differences between the COVID and non-COVID epoch. However, with very large numbers, small differences are ‘significant’ in a statistical sense and there is no pattern in the data, sometimes favouring the pre-COVID era (emergencies and elective surgery) and sometimes favouring the COVID era (cancer).

There was no evidence in the data that people from ethnic minorities or from lower socio-economic groups experienced a greater drop in access compared to white or higher socio-economic groups. Over most operation types, the proportions of cases classified as ‘white’ declined in favour of those classified as ‘unknown’.

### Strengths and limitations

This is the first comprehensive study using a national dataset of all hospital admissions in England examining disruptions in a number of areas including routine, complex, emergency and elective procedures. Previous studies have been limited by examining disruptions in surgical care of people from single centres or regions and mainly related to cancers,^14^ patient experiences,^15^ and single routine or emergency procedures.^16-18^

Our study is limited to a particular range of surgical procedures because we wanted to see how the system had been able to cope with procedures of different types. However, these types do not serve as a classification system for two reasons. First, they overlap. For example, cancer operations are urgent because they are time critical. Second, they are hard to define or categorise in much the same way as a tree and a bush. Thus, our criteria simply illustrate features of different surgeries that act as barriers (e.g. complexity) or facilitators (e.g. degree of urgency) to access to surgery during an acute rise in hospital admissions. We think our findings show that when an operation is complex, not just technically but in terms of the pathway patients follow, then this acts as a barrier even if surgery is time-critical. This study does not seek to identify where on the patient pathway barriers occur databases.

### Implications

While reductions in surgery were attenuated in the second peak compared to the first peak, it would be unwise to assume that this trend would continue in a third peak of similar intensity. Policy in the face of rising admissions must consider this risk.

As stated above, our study does not identify where barriers occurred. While in previous studies we have studied ambulance admissions and hospital treatment of stroke and myocardial infarction, a more elegant solution would be to link these and other sources of data.

There are many different ways these data can be interpreted in terms of policy beyond this particular pandemic. One interpretation is that the service coped well given the extraordinary pressure it was under, and it would be wasteful to maintain sufficient capacity to weather future epidemics on a similar scale. Another might be that the health loss from not being able to treat early cancer and maintain transplant services is too large to ignore and some sort of contingency planning should be a priority for the future as we have argued in relation to the nuclear threat.19 We hope that the data presented here will usefully inform this debate.

## Supporting information

STROBE Checklist

## Data Availability

Linked HES and ONS data may be obtained from NHS Digital via the Data Access Request Service (DARS) and are not publicly available.

## GUARANTOR INFORMATION

The corresponding author attests that all listed authors meet authorship criteria and that no others meeting the criteria have been omitted.

## STATEMENT ON PATIENT INVOLVEMENT

No patients were involved in setting the research question or the outcome measures, nor were they involved in developing plans for recruitment, design or implementation of the study. No patients were asked to give advice on interpretation or writing up of results. There are no plans to disseminate the results of the research to study participants or the relevant patient community.

## TRANSPARENCY DECLARATION

The lead author affirms that this manuscript is an honest, accurate, and transparent account of the study being reported; that no important aspects of the study have been omitted; and that any discrepancies from the study as planned and registered have been explained.

## ETHICS APPROVAL

This observational study was registered with the local Clinical Audit Department (Clinical Audit Registration and Management System number 16961). The University of Birmingham Science, Technology, Engineering and Mathematics Ethical Review Committee board ruled that ethical approval could be waived and patient consent was not needed. Data were used in line with the data sharing agreement with NHS Digital.

## DISCLAIMER

Views expressed are those of the authors and not necessarily those of the NIHR, the NHS or the Department of Health and Social Care.

